# Perceptions on undertaking regular asymptomatic self-testing for COVID-19 using lateral flow tests: A qualitative study of university students and staff

**DOI:** 10.1101/2021.03.26.21254337

**Authors:** Marta Wanat, Mary Logan, Jennifer Hirst, Charles Vicary, Joseph Jonathan Lee, Rafael Perera, Irene Tracey, Gordon Duff, Peter Tuffano, Thomas R. Fanshawe, Lazaro Mwandigha, Brian D. Nicholson, Sarah Tonkin-Crine, Richard Hobbs

**Affiliations:** Nuffield Department of Primary Care Health Science, University of Oxford; Nuffield Department of Primary Care Health Science, University of Oxford; National Institute for Health Research (NIHR) Oxford Biomedical Research Centre, University of Oxford Hospitals NHS Foundation Trust; Merton College, University of Oxford; St Hildas College, University of Oxford; Said Business School, University of Oxford; Nuffield Department of Primary Care Health Sciences, University of Oxford

## Abstract

**Background:** There has been an increased interest from governments in implementing mass testing for COVID-19 of asymptomatic individuals using Lateral Flow Tests (LFTs). Successful implementation of such programmes depends on several factors, including feasibility, acceptability and how people act on test results. There is a paucity of studies examining these issues.

**Objective:** We aimed to examine experiences of university students and staff with experience of regular asymptomatic self-testing using LFTs, and their subsequent behaviours.

**Methods:** We invited people who were participating in a ‘weekly testing’ feasibility study. We conducted semi-structured remote interviews between December 2020 and January 2021. Additional qualitative data from a survey were also analysed. Data were analysed thematically.

**Results:** We interviewed 18 and surveyed 214 participants. Participants were motivated to regularly self-test as they wanted to know whether or not they were infected with SARS-CoV-2. Most reported that a negative test result did not change their behaviour but it did provide them with reassurance to engage with permitted activities. In contrast, some participants reported making decisions about visiting other people when they would not have done so otherwise, because they felt reassured by a negative test result. Participants valued the test training but some participants still doubted their ability to carry out the test. Participants were concerned about safety of attending test sites with lots of people and reported home testing was most convenient.

**Conclusions:** If governments want to increase uptake of LFT use, clear messages highlighting the benefits of regular testing for family, friends and society in identifying asymptomatic cases are needed. This should be coupled with transparent communication about accuracy of LFTs and how to act on either a positive or negative result. Concerns about safety, convenience of testing, and ability to do tests need to be addressed to ensure successful scaling up asymptomatic testing.

## Introduction

Infection prevention and public health strategies rely on early detection of disease to prevent spread^1^ . So far in the COVID-19 pandemic, the UK government has deployed strategies such as various grades of restriction in population movement, required social distancing, use of face coverings in public, and testing for patients with typical symptoms using laboratory COVID-19 polymerase chain reaction (PCR) tests. While these tests are considered the gold standard for diagnosis, they have limitations, including slow turn-around time, specialist facilities needed for processing, detection of non-infectious SARS-CoV-2 particles, limited access, and high costs^2^.

Recent data showing that 1 in 3 people infected with SARS-CoV-2 may not show symptoms, prompted an increased interest from governments in the feasibility of testing asymptomatic individuals using lateral flow tests (LFT) ^3^ . In fact, in a number of countries^4 5^, including the UK^6^, these devices are part of government strategies for easing of lockdowns. The Liverpool Testing Programme was one of the first to examine, alongside the accuracy of LFTs, the feasibility and acceptability of such programmes in an area with high COVID-19 prevalence^7^. University campuses were also identified as potential sites of high COVID-19 transmission^8^, which resulted in pilot studies in a number of universities testing asymptomatic populations^9-11^.

Asymptomatic testing (screening) has attracted a lot of attention, with many highlighting that LFTs can give people false reassurance^12^, and others stressing that targeted testing could help reduce transmission and be part of the lockdown exit strategy^14^. As there are no clinical trials of SARS-CoV-2 screening, there is uncertainty of the effect. The supposition is that identifying positive cases willing to go on to self-isolate rapidly and cheaply could reduce infections more than any increase in infections from falsely reassuring people with false negative results. Furthermore, repeat testing with LFTs for SARS-CoV-2 infections, if shown to be feasible, will markedly improved the relatively poor sensitivity of the tests. For any given test, people’s behaviours will determine this balance. Screening pilots usually request that participants do not change their behaviour as a result of being tested and receiving a negative result. This is also an assumption of modelling studies that have proposed this strategy. The evidence from point-of-care tests for other conditions is that tests are part of complex interventions that change behaviours in unpredictable ways^15^. Evidence is therefore needed on participant perceptions on the use of COVID-19 LFTs.

Successful implementation of asymptomatic testing using LFTs depends on a number of factors, including whether tests are acceptable and feasible to conduct. It is also important to know how people interpret a test result as this may have implications for how they act on it. These issues could be understood within Protection Motivation Theory (PMT)^16^. This theory frames the intended response to a potential health threat as being based on two factors: i) threat appraisal and ii) coping appraisal. Threat appraisal is determined by two components: the perceived severity of the threat and perceived vulnerability to the threat. In the context of COVID-19, these may be seen as how likely a person thinks they are to become infected with the virus and how serious a person perceives the consequences of being infected (e.g. being admitted to hospital, infecting others etc.). This then influences how people may view the benefits of implementing protective behaviours. Coping appraisal is affected by three components: perceived efficacy of one’s responses to the threat, associated costs, and self-efficacy. Again, in the context of COVID-19, this may be understood as to what extent one thinks that protective behaviours (e.g. wearing face masks, social distancing, etc.) are effective, what the costs associated with these protective behaviours are, and how confident one feels in carrying out these behaviours. From the PMT perspective, a LFT result may influence coping appraisal and beliefs about how well people feel their engagement with protective behaviours are working. This may have consequences for one’s behaviour. For example, a positive LFT result may indicate to people that protective behaviours they have carried out to date are unlikely to have been insufficient and they may motivate others to follow guidance more strictly. Alternatively, a negative test result may increase people’s beliefs that the protective behaviours they are carrying out are sufficient. This may motivate them to continue with their current behaviour or potentially lead them to relax some of their efforts.

Few studies have examined acceptability and feasibility of asymptomatic testing in the community^7 11^ and even fewer have focussed on these issues in relation to regular testing (rather than one-off) ^9^. Previous studies have focussed on PCR^9 17^, rather than LFT, testing. People’s views and understanding of the accuracy of such tests have also not been explored. Our study aimed to address this gap by examining experiences of university students and staff of regular self-testing using LFTs with the aim of identifying key lessons for future asymptomatic testing programmes.

## Methods

### Study recruitment

The ‘Feasibility and Acceptability of community COVID-19 Testing Strategies’ (FACTS) study was a mixed methods cohort study conducted at the [X]. It was approved by the [X] in October 2020. X University students and staff were invited to participate in the study through a number of departments via an email invitation, providing information on the study including a participant information sheet (PIS). Participants who agreed to participate were required to download the ‘CoronaVirus health Monitor (CVm)-Health’ app developed by X. This bespoke app recorded consent to the study, symptoms, test results, an upload of a photograph of the completed test, and responses to a one-item acceptability question. Participants were invited to a training session before undertaking weekly testing.

### LFT training

The email invitation also contained a 5-minute video explaining how to prepare to test, perform the swab, extract the sample, test the sample and read the results. The study used the Innova Rapid SARS-CoV-2 Antigen Test Kit (Innova Medical Group, US) ^18^. Participants received either face-to-face or online training. Details of training are reported elsewhere^19^. Face-to-face training was carried out at participants’ x [anonymised place] or department. Participants were talked through the aims of the study, followed by a demonstration of the swab, sample preparation, recording and interpreting the test result. Following the demonstration, participants performed the test, with support from the trainers if required. When 30 minutes had elapsed since applying the sample to the test device, participants were asked to interpret the result, record this on the app, and photograph the result using the app. The trainers visually checked each test result and confirmed whether the participant’s interpretation of the result was correct. For a small number of participants, live on-line training was offered to participants who collected test kits in advance. This was delivered by two trainers via Zoom and involved similar procedures to the face-to-face training. The session was interactive so participants could ask questions.

The PIS informed participants that rapid tests are not as reliable as, or a replacement for, a PCR test. In the event of a positive test result, participants were advised to self-isolate and book a PCR test through the University COVID-19 testing service; in the event of a negative test, participants were advised to follow normal infection prevention procedures. This information was repeated during the training where participants were also told that there is limited evidence on the accuracy of the test in asymptomatic people. At the time of the study the Innova test had not received MHRA authorisation for asymptomatic testing, nor for self-testing, and the test use was under research ethics.

While the original plan was to provide all participants with the testing kits to take home, this was not possible in the initial stages of the study. That meant that participants who joined early in the study had to attend their work or study site for testing for several weeks prior to the ‘take-home’ kit being available.

### Interview recruitment

At the time of study enrolment, participants were asked whether they would be interested in being contacted about taking part in an interview on their experiences of testing. A selection of participants who agreed were invited, using purposive sampling, in order to obtain variation in university role (student or staff) and department/[anonymised place]

### Data collection

A semi-structured interview guide was developed based on the primary research questions and informed by the existing literature on COVID-19 testing in order to elicit key determinants of behaviour (Appendix 1). Participants were asked about their views and experiences of using the tests, their reasons for taking part in the study, barriers and facilitators to undertaking regular testing, trust in test results and intentions to act on a positive result. After obtaining consent, interviews were conducted over the telephone or online by an experienced qualitative researcher and audio recorded. Interviews continued until data indicated saturation^20^. We conducted rapid data collection and analysis concurrently.

As part of the wider study, we also conducted a survey examining participants’ views of regular testing. The survey consisted of 13 questions (Appendix 2) and a free text comment box. The survey was sent to all study participants via email. Based on the free text comments, we created an initial framework consisting of nine categories that captured key areas of interest. Using the framework, detailed summaries of interview data, including verbatim quotes, were made directly from the audio recording after each interview^21 22^. These were further changed and then used to create themes and sub-themes. Throughout the process, the qualitative researchers (MW and STC) met regularly to identify key patterns in the data and to compare and contrast emerging findings. This method is deemed a pragmatic and efficient approach to collect and analyse data rapidly during a public health emergency^21 22^.

## Results

734 participants across a number of departments and colleges took part in the study (October 2020 to January 2021) and performed 3187 LFT tests. The study covered a period of various restrictions, at first Tier 2 restrictions, followed by full lockdown with an exception of household mixing allowed for one day at Christmas. Participants completed a mean of 4.3 tests over a mean of 4.8 weeks, but these varied by [anonymised place] or department^19^.

431 of 733 (59%) participants indicated on the consent form that they would be interested in taking part in an interview. Fifty-two were approached and 18 interviews were conducted (response rate 35%). Of these, 3 were undergraduate students, 3 were postgraduate students and 12 were staff. Each interview participant had completed between 3 and 10 tests during the whole study period (mean 7.7). None of the interviewed participants received a positive LFT result during the study. The interviews took place between 11^th^ December 2020 and 18^th^ January 2021 and lasted between 17 and 43 minutes (mean 26 minutes). In addition, 214 participants completed the survey (29%); 62 provided additional free text comments. 61 (29%) were undergraduate students, 81 (38%) were postgraduate students and 72 (33%) were staff. Each survey participant completed between 1 and 13 tests (mean=5.8).

We identified four themes, which we report below with supporting quotes.

### Theme 1: Perceived benefits to regular testing

Participants reported three main benefits of taking part in the study and having access to regular self-testing. Firstly, they wanted to check regularly whether or not they were infected with SARS-CoV-2, to reduce their fear of unknowingly infecting others, which was a concern they mentioned frequently. For some participants this extra knowledge was related to their perceived risk of becoming infected, and access to regular testing was particularly important for reassurance.

> *I live in a densely populated area so I am exposed to risk a lot. When I began testing, I was regularly travelling by train […] so I kept at it because I was in a relatively risky situation. [P15, Staff, Interview]*

In addition, participants wanted to know if they were infected so they could take appropriate action, i.e. self-isolate and thus minimise the risk of spreading the virus.

Secondly, some students highlighted that deciding to self-test with a LFT was perceived as a personal choice and therefore more acceptable than undertaking NHS or university testing when experiencing symptoms. The university protocol for symptomatic testing required everyone in the household to enter into isolation at the time of getting a test rather than at the time of getting a positive result. As participants explained, peer pressure may prevent people from doing NHS or university testing.

> *Getting an NHS test is such an ordeal and in a university context, there is pressure not to get tested because getting that test puts your whole house into a lockdown. This test removes barriers […] You do it as a personal choice and not something where you get permission from the whole household to get tested [P2, Student, Interview]*

Finally, all interviewees wanted to support COVID-19 research to contribute to fighting the pandemic. Some hoped that the study would provide evidence for the value of asymptomatic testing, which if implemented widely, would help ease the restrictions.

### Theme 2: Perceptions of test accuracy and its implications

#### Sub-theme: Knowledge and views of test accuracy

While most of the participants perceived the test as providing reassurance about whether they were infected, views on test accuracy varied.

Overall, participants mostly accepted that tests were not 100% accurate. They saw them as just one of the measures to try to avoid spreading the virus (among social distancing, face masks and vaccines in the future). Some participants sought their own information on the accuracy of LFTs in general or had heard information from family and friends. The perceived accuracy varied greatly, with participants citing figures between 50 and 90%. It is important to highlight that often the same figure was seen as reassuring by some participants, and less so by others.

> *I am sceptical because someone who works in the industry told me that some hospital stopped using the tests because with poor training it has an effectiveness rate of 50% and although I am fairly confident that I am doing everything correctly, I am being more cautious [P19, Student, Interview]*
>
> *I talked to a friend who is a nurse; and she said that they are around 60% which is a decent percentage to be accurate [P18, Staff, Interview]*

Some participants lacked any recalled information on accuracy of the tests.

> *Whilst difficult to comment on accuracy, as this info has not been given, I am happy with administering the test [P25, Staff, Survey]*

#### Sub-theme: The implications of knowledge on one’s behaviour

Participants’ views on test accuracy were important when making decisions about their behaviour. Participants did not view a negative test result as permission to break government guidelines, but reported that negative tests increased their confidence to engage with activities that were allowed.

> *Some time ago, we made a decision to visit one set of relatives for Christmas. […] I am not sure [the test result] changed our behaviour but it reassured us that I am going to have two tests during that time and if they are both negative that gives you a bit of reassurance that this is a reasonable thing to do [P11, Staff, Interview]*

Crucially, some participants did make decisions, based on negative test results, about engaging with activities where there was potential for transmission (for example seeing a relative or extent of contact with relatives at home) because they were unaware about tests not being 100% accurate. When later learning that tests were not 100% accurate, participants were concerned about their decisions.

> *I have read online about the reliability of the tests and initially that gave me a lot of confidence as my relative lives near, who is 88, and when I had a negative test I felt that I could go and have a cup of tea with her and then I read that the reliability was, that there were a lot of false negatives, so some of the figures were down in the 50s or 60s, so 60% and then you thought ‘oh this is not that reassuring if it is only 60%’ [P14, Staff, Interview]*
>
> *The level of accuracy of the LFT tests (and in particular that they miss a very significant amount of positive COVID carriers) was not communicated to me, so I think this gave me a false sense of security that I did not have COVID. [P59, Student, Survey]*
>
> *I sorted my Christmas arrangement so that I was going home the day I took the test so that has then given me a peace of mind. [P17, Student, Interview]*
>
> *I have done all these tests which were negative and after the 3rd test I was less careful for sure [P5, Student, Interview]*
>
> Finally, some participants were unsure whether the information they had read about LFTs was relevant to the test they had been using. They highlighted the difficulty of making a decision on whether to engage with certain (allowed) activities or not.
>
> *I read in the newspapers that when done by trained medical staff the tests are only 75% accurate, and by non-medical staff 50%. I don’t know if I understood this information correctly but that is my worry. […] So if I have 50-50 success rate is that a good thing or is it better not to know and just assume that you are not clear and act accordingly or does this negative reading gives you a license to be more, well for example to see your family at Christmas. And I read that it does not gives me a license to see my mum and dad. [P3, Staff, Interview]*

Overall, participants were aware of (the small risk of) false positives but reported that they ‘would trust a positive test result completely’, prompting them to self-isolate immediately and seek additional RT-PCR testing.

### Theme 3: Extent of confidence in ability to do the tests

The majority of participants felt that the training they received enabled them to feel confident about doing the tests. Watching the video followed by a face-to-face interactive training session was reported to be beneficial. In contrast, any discrepancies between what was in the video and the face-to-face training, for example in relation to how to prepare the test (which was due to the fact that video used was prepared for a different university), was confusing and, at times, decreased confidence in the accuracy of the test.

While doing tests repeatedly increased participants’ confidence, a number of participants were still unsure whether they were doing the test correctly, especially the tonsil swab. Some questioned whether an incorrect swab would make test less reliable.

> *I have very strong gag reflex so I am unable to reach my tonsils. I was told by the nurse: ‘just try your best’, which was not helpful. I have not been able to get an answer on whether it is important to swab the tonsils and whether it is a big problem or not. [P8, Staff, Interview]*

Participants who were unsure where the tonsils were suggested that a more realistic picture of tonsils (rather than a cartoon) might be helpful.

Doing the tests at home was easier as participants had access to mirrors rather than having to rely on their phone cameras to do the test on site. When doing tests at home, having a card which summarised the instructions was also suggested, as instead participants had to re-watch the video every time they were unsure about some aspect of the self-testing. In contrast, doing tests on site was perceived as helpful by some participants as they could ask other participants for tips and check whether they “were doing it right”, which they found reassuring. Also, seeing other people experiencing physical sensations such as watering eyes as a result of doing the test, was helpful in knowing that this is what one can expect. However, test preparation on site was difficult as participants reported having to prepare the tests on small tables.

### Theme 4: Barriers and facilitators to regular testing

All interviewees experienced swabbing as uncomfortable, at least to a certain extent, with some reporting having a strong gag reflex and testing causing sneezing or watering eyes. However, most participants highlighted that these sensations were temporary, manageable and did not decrease their motivation to continue self-testing and were a “small price to pay” for getting reassurance on whether they were infected (as described in theme 1).

Participants who were able to take a number of testing kits home seemed to see testing as relatively easy to fit tests into their weekly routine. In contrast, for participants who did not get packs to take home and who had to go to their [anonymised place] or department to self-test, it was an inconvenience and caused frustration, especially as testing took place over several weeks. This was especially the case for staff.

While training in a group was perceived as beneficial (as described in theme 2), some participants were also concerned about the safety of getting tested on site, around other people, especially if they had not been going out much.

> *Also, though this has now been resolved, it was quite frustrating that we were expected to conduct the test in person in a lecture hall with many other students for the first few weeks, as this was the biggest personal risk I took each week in terms of COVID exposure [P10, Student, Survey]*.

Some staff participants found the experience of self-testing in a group awkward and embarrassing when being with other colleagues. Others commented that being with people they knew provided a nice sense of camaraderie if “they were all having a laugh”. Finally, participants wanted a reminder to do the test when it was due each week, and some felt this could provide additional encouragement.

## Discussion

We found that interviewees were motivated to conduct regular testing as they wanted to know whether or not they were infected with SARS-CoV-2. While most participants accepted that the test was not 100% accurate, many could not quantify this further and estimates of test accuracy varied greatly among participants. Although information on test reliability was provided in the group and online training sessions, this was not recalled by all. Importantly, most reported that a negative test result did not change their behaviour but it did provide them with reassurance to engage with permitted activities. However, there were exceptions, with some participants who reported making decisions about contact with other people when they would not have done otherwise, because they felt reassured by a negative test result. Participants valued the training but some participants still doubted their ability to do the test, which in turn decreased their confidence in accuracy of test results. Participants also raised the importance of safety and convenience when attending for tests on site.

### Comparison with existing literature

Participants in our study wanted to have regular testing to reduce their fear of accidentally infecting their family, friends or other people in their community, while also wanting to contribute to fighting the pandemic. This is in line with the Liverpool COVID-SMART study, which found that people signed up to have a test as they wanted to protect their families, friends as well as local hospitals and NHS workers and saw taking part as “the right thing to do” ^7^. Only one study in a university setting explored these issues, albeit involving RT-PCR tests, and also found that “helping to keep the campus safe”, “contributing to the national effort to control the virus” and “being involved in COVID-19 research” were the top three reasons for taking part^17^. Further analysis also revealed that the number of returned tests was positively correlated with increased worry about friends and family contracting COVID-19^17^. Our study also highlights the importance of the perceived benefits but in the context of regular rather than one-off testing and using LFTs. It also suggests that asymptomatic testing using LFTs may be perceived as more accessible and acceptable for students, in comparison to NHS or University testing, which has not been identified before.

Importantly, our study found that while most participants understood that the test was “not 100% accurate”, estimates of test accuracy varied greatly among participants. Most reported that negative test results did not change their behaviour but it did provide them with reassurance to engage with permitted activities, which they had planned to participate in. However, some participants felt reassured by the test and reported making decisions involving contact with other people, when they would not have done otherwise. Previous studies have only explored these issues for antibody testing^23^. The survey conducted as part of the Liverpool COVID-SMART study indicated that some participants had concerns about test accuracy^7^ and one study in a university setting found that 79.6% of participants were confident in the outcome of their PCR test^12^. Our study highlights that people’s understanding of the extent to which LFTs are accurate varied, with potential implications for their behaviour.

It is important to note that no participants who answered the survey or who took part in an interview tested positive for COVID-19 during the study. Participants indicated that they would trust a positive result and self-isolate as advised although this may reflect socially desirable responses. It is notable that none of the participants indicated they would have any difficulty in self-isolating if necessary, which is positive for any future roll out in universities but again reflects that this population is not representative of the general population, where only around 20% of symptomatic people might complete self-isolation^24^.

Students reported negative peer pressure when wanting to engage NHS or university testing when experiencing symptoms as this would result in those peers being forced to self-isolate. Having access to the LFTs removed these barriers for some, allowing them to the personal choice to engage with testing. Therefore when considering a mass testing system it is important to take into account that one process may not be effective in reaching all those who would otherwise be willing to engage.

Finally, while our participants described swabbing as being uncomfortable, they felt that the perceived benefits outweighed the burden of doing the tests. Having access to a number of tests which they could do at home made it easier for participants to take part, while doing the testing on site provided an opportunity for feedback on how well participants were doing the test but magnified safety concerns. Misinformation related to perception of the risk of infection at test sites, and the need to have physical contact with centre staff, have been described before^17^.

The study findings can be partially explained by Protection Motivation Theory^16^. Qualitative data from this study highlight that from the PMT perspective, a negative test reassured people that the protective behaviours they had engaged with previously had been effective. Regular testing meant interviewees felt they had regular feedback about whether they were infected. They felt that the costs (such as time, physical sensations) of undertaking the test were small and mostly felt confident in conducting the test. For a minority however, lack of information or misunderstandings related to the accuracy of the test seemed to affect their coping appraisal as it gave them more confidence that they were not infected, which lead them to engage with additional activities, which they had previously avoided, involving contact with other people.

### Strengths and limitations

This is the first qualitative study examining views and experiences of students and staff of regular asymptomatic SARS-CoV-2 testing in a university setting using LFTs. It highlights a number of key issues related to acceptability and feasibility of regular testing as well as its behavioural implications. We note some limitations. The mean number of tests conducted by each interview and survey participant was higher than the mean number of tests in non-interviewed participants, so our sample may over-represent those who continued to test regularly. Additionally, the FACTS participants were university student and staff volunteers, whose motivation to participate and perceived benefits may be different from those in the wider university population, and other non-university settings. The majority of interview participants were also staff. We adopted rapid qualitative analysis to aid identification of key issues but full transcription of qualitative data could have minimised the potential for errors of interpretation. However, we discussed interpretation of data on a regular basis with other members of the team and extensive notes have been made after each interview.

### Implications for policy and practice

Our study indicates that messages highlighting the benefits for family, friends and society in identifying asymptomatic cases and contributing to fighting the pandemic and ultimately lifting lockdowns might be beneficial for encouraging regular use of LFTs. However, these need to be coupled with clear and transparent communication about accuracy of LFTs. In context diagnostic accuracy studies are therefore a pre-requisite.

Information about accuracy of tests is important but given that the same reported accuracy of the test might be perceived by different people as more or less favourable, it is crucial that this is framed within clear messages on what it means for an individual’s behaviour (i.e. the need to follow COVID-19 safety measures). This is especially important for testing in workplaces or schools (as currently planned in the UK^6^) where a negative test may allow people to return to their study or workplace and will consequently involve contact with other people. Advice that supports people to continue physical distancing, hand hygiene and mask wearing in the context of a negative test is crucial. The timing of providing this information is important, as well as the need to provide clear information about test accuracy in advance, and repeated, to prevent people creating a mental model which is then difficult to change^25^. This may also be relevant in the context of more extensive vaccine rollout, as some have raised concerns that vaccination may encourage people to ignore public health messages^26^. Recent reports of implementation of asymptomatic testing in local authorities in England showed limited communication about these issues. A study involving a search of local authority websites found that 47% did not explain the limitations of LFTs or that people should continue following safety measures and restrictions despite a negative result and highlighted a lack of standard messaging on test accuracy^27^.

When scaling up regular asymptomatic testing, it is important to also consider potential concerns about convenience of testing, and people’s confidence and ability to do the testing. In settings where people may be tested on site, safety and convenience may be important to consider. Concerns about physical sensations also need to be addressed. Whereas, for those who are sent tests to take at home, clear information on testing procedures and a reminder to take the test will be of importance. We also found that training materials provided should mimic what is delivered to participants face-to-face, as when trying to learn about and understand how to use these new devices any deviation in the two can lead to confusion and uncertainty.

## Conclusions

University students and staff found access to regular testing using LFTs beneficial. Importantly, in contrast to how LFTs are mainly being used in the UK, self-testing proved feasible and acceptable and, further, that the ability to self-test at home rather than in a public space was welcomed. This may explain why the uptake of testing in this pilot was much higher than the rates observed in the standard UK model of self-swabbing but staff testing of the LFT in public testing venues. Diagnostic information about testing needs to contain clear and transparent communication on test accuracy, and the need to adhere infection prevention and transmission measures in the event of a negative test. The findings provide an insight into key barriers and facilitators around safety, convenience and confidence to self-test which need to be addressed for successful implementation and scaling of asymptomatic testing using LFTs.

## Data Availability

 The data are not publicly available due to their containing information that could compromise the privacy of research participants.

